# Evaluation of a training course for general practitioners within the Melanoma Multimedia Education Programme of the Italian Melanoma Intergroup: study protocol

**DOI:** 10.1101/2023.12.27.23300576

**Authors:** Ignazio Stanganelli, Serena Magi, Lauro Bucchi, Emanuele Crocetti, Silvia Mancini, Rosa Vattiato, Stefano Falcinelli, Patrizia Re, Davide Melandri, Marco Brusasco, Sara Gandini, Fabio Falcini, Federica Zamagni, the FAD MelaMEd Working Group

**Author notes:** **Corresponding Author:** Federica Zamagni, Emilia-Romagna Cancer Registry, Romagna Cancer Institute, IRCCS Istituto Romagnolo per lo Studio dei Tumori (IRST) “Dino Amadori”, via Piero Maroncelli 40, 47014, Meldola (FC), Italy.

## Abstract

The text discusses the role of general practitioners (GPs) in the prevention and early diagnosis of melanoma, a type of skin cancer. It highlights the need for GPs to be able to recognize suspicious skin lesions and refer patients to specialist dermatology centers. However, many GPs lack comprehensive training in diagnosing melanoma. The text mentions that various training courses have been conducted for GPs, but their impact on clinical practice has been limited.

The “MelaMEd Programme” is an e-learning course developed by the Italian Melanoma Intergroup (IMI). The program aims to provide GPs with comprehensive knowledge of melanoma prevention, diagnosis, and treatment. It includes an e-learning section and a dedicated platform called MelaMEd platform, which offers a multimedia atlas of melanoma.

The objective of the study is to evaluate the impact of the MelaMEd programme on GPs’ diagnostic accuracy, knowledge of melanoma, and management of suspicious lesions. The methodology involves administering pre-training and post-training questionnaires to participants, assessing their diagnostic skills and evaluating the training course’s effectiveness.

The study aims to demonstrate the effectiveness of the MelaMEd programme in improving GPs’ ability to recognize and manage melanoma. It also seeks to identify areas for improvement and recommend interventions to enhance diagnostic accuracy. The results will be analyzed statistically using descriptive, univariate, and multivariate analysis methods.

## 1. RATIONALE AND BACKGROUND INFORMATION

### 1.1 The role of general practitioners in the prevention of melanoma

Several public information and education initiatives on self-skin surveillance have been carried out in many European nations throughout the years. Although very confusing, these initiatives have shown to significantly improve the time to diagnosis and survival from cutaneous melanoma.^1^

The primary prevention for skin malignancies, the clinical recognition of “suspicious melanoma” and the identification of patients at risk of melanoma are the recommended strategies by Europe’s Beating Cancer Plan^2^ and by Italian and international guidelines.^3–6^

According to these recommendations, general practitioners (GPs) and also paediatricians play an active role in the referral pathway to the dermatology center. Being the natural “link” between patients and the specialist reference centers, GPs must be able to recognize the skin lesions with clinical-morphological characteristics that are consistent with early-stage melanoma. Additionally, they must be at least somewhat familiar with the entire fundamental melanoma diagnostic-therapeutic pathway. In other words, their role is mainly to select to high-risk population and perform a preliminary differential diagnosis, i.e., to select patients susceptible to specialist diagnostic investigation. Nonetheless, many GPs lack of comprehensive training in diagnosis, patient selection, and the diagnostic-therapeutic pathway for skin malignancies, especially melanoma.

A recent review has collected the results of training courses for GPs on melanoma prevention. Interventions were heterogeneous and varied widely in the design and teaching methods, with live or online courses lasting from 5 minutes to 24 months.^7^ Whilst several initiatives have demonstrated significant improvements in melanoma knowledge and expertise, only a few studies have revealed positive changes in clinical practice through histological review.^7^

The adequate triage of skin lesions and the knowledge of the essential basis of the melanoma diagnostic-therapeutic pathway allow an effective and efficient cooperation between GPs and specialist structures. According to the Italian Oncology Plan 2023-2027, the training of healthcare workers is considered “one of the best investments to guarantee high levels of performance” and “highly dynamic training interventions are recommended because they concern a continuously evolving sector with regards to care models, technological innovations and the indispensable aspects of humanization and respect for the person”.^8^ For these reasons, the importance of multimedia technologies in education has been highlighted and the widespread adoption of multimedia e-learning tools has been recommended. In Italy, the effect of formal training on GP has been seldom evaluated and the impact of multimedia educational programme has never been investigated.^9,10^

### 1.2 The “MelaMEd Programme” and the evaluation of its training impact

The Melanoma Multimedia Education (MelaMEd programme), one of the projects developed by the Italian Melanoma Intergroup (IMI), aims to provide physicians, mainly general practitioners, with a comprehensive understanding of the primary and secondary prevention of cutaneous melanoma and a broad overview of diagnostic and therapeutic procedures. The MelaMEd programme is hosted on a dedicated platform: an educational area which includes an e-learning section, with training courses accrediting Continuing Medical Education (CME) points.^11^

This multimedia training course is aimed at building up, at a local level, a network between multidisciplinary teams dealing with melanoma and healthcare workers. In particular, this training course has been recommended to GPs, healthcare workers who are part of multidisciplinary melanoma teams and those dealing, at a local level, with the management of patients with melanoma.

The MelaMEd programme provides the knowledge and experience that will enable participants to recognize skin lesions requiring specialist dermatological consultation by applying validated methods. Recent scientific evidence on the association between the trend of annual dermatology outpatient visit rates, skin biopsy rates and melanoma incidence rates has demonstrated a limited selection of the population at risk and of suspicious melanoma by GPs.^12^ This epidemiological data undoubtedly represents an important basis for the implementation of specific training courses in the management of the diagnostic and therapeutic pathway of melanoma. These premises also highlight the need to carry out an evaluation of the training course and represent the rationale for this protocol.

## 2. OBJECTIVE

In the field of prevention, the expression *demonstration project* refers to projects in which a preventive tool of proven or accepted effectiveness (a dietary correction, a test for early diagnosis, etc.) is used in practice with the aim to “show something and explain how it works”.^13,14^

The evaluation of these projects is essentially descriptive, and the MelaMEd programme has similar characteristics. We plan to provide a teaching activity on a large scale to a potentially very large number of subjects, i.e. in a real rather than experimental situation.

To establish if and how this preventive strategy works, we have identified as a primary objective the MelaMEd programme impact evaluation in terms of improvement in 1) diagnostic accuracy (recognition of suspicious lesion), 2) basic knowledge of the disease, and 3) management of suspicious lesion.

Our secondary objective is to gather useful information about how to enhance diagnostic accuracy and fundamental knowledge of melanoma. To address the first point, we will identify the areas in which the basic knowledge of melanoma needs to be strengthened. To address the second point, we will assess the factors that may influence the accuracy of the diagnosis and recommend appropriate intervention.

## 3. STUDY DESIGN

### 3.1 The MelaMEd programme

The MelaMEd programme consists of a dedicated asynchronous e-learning course entitled “Early diagnosis and management of the therapeutic diagnostic pathway of melanoma”. The asynchronous e-learning course includes, among the educational contents, a link to the MelaMEd platform, a website developed by IMI devoted to melanoma diagnosis and treatment. Direct access to the MelaMEd platform is possible from the asynchronous e-learning course.

The asynchronous e-learning course is free of charge and is delivered on behalf of the Federazione Nazionale degli Ordini dei Medici Chirurghi e degli Odontoiatri (FNOMCeO), a national association including all the Italian community of physician and dentists. The asynchronous e-learning course was activated on June 1st, 2022 and available until December 31, 2023.

The asynchronous e-learning course has included video presentations, with audio and visual information on following topics: identification of risk groups; sun exposure pattern, photoprotection, melanoma diagnosis using the mainly clinical Prediction Rules as ABCDE,^15,16^ EFG^17^ and the “Ugly Duckling” sign,^18,19^ histopathological diagnosis, surgical and medical therapy and finally interactive clinical cases in the various spectrums of melanoma progression with the related decision-making points.

For each topic, MelaMEd programme has an interactive function which allows the user to further explore any aspect of the course material available in ‘switching’ modality with the MelaMEd platform.

The MelaMEd programme is a real revolution in the panorama of multimedia medical training, offering interaction and integration functions connecting the asynchronous e-learning format and the MelaMed platform.

### 3.2 The MelaMEd platform

The MelaMEd platform^11^ is a vast virtual library, a multimedia atlas, free of charge, quick to access and easy to consult. Divided into fundamental chapters containing iconographic documentation of the clinical, dermoscopic and histological aspects of melanocytic pathology, the site is geared to raising awareness of the importance of identifying melanoma at an early stage. The MelaMEd platform also details surgical and medical treatments, stage and prognosis according to the latest guidelines accredited by Italian National Health Authority and by other European and international organizations.^3–5^

More specifically, the MelaMEd platform is divided into 10 chapters: epidemiology, risk factors, with particular attention to ultraviolet radiation (chapters 1,2 and 3), clinical and differential diagnosis of melanoma facilitated by the use of non-invasive cutaneous techniques (chapters 4, 5, 6 and 7), histological diagnosis, medical and surgical therapy, staging, prognosis (chapters 8 and 9) and finally the most frequently asked questions from patients, with related answers (chapter 10). Each multimedia chapter includes a combination of more than one media type such as text, symbols, pictures, audio, video, with the use of static and dynamic images, as well as links to the most recent literature and the most updated official websites. The MelaMEd platform’s contents and descriptions are listed in Table 1.

**Table 1.** Number, name and description of the chapters included in the MelaMEd platform.

| Number | Name | Description |
| --- | --- | --- |
| 1 | Epidemiology | Epidemiology of melanoma in terms of incidence, mortality and survival that reports the main references at international and national level |
| 2 | Risk factors | Comprehensive review of following mainly risk features: genetic, phototypes, melanocytic nevi, ABCDE and EFG rules, ugly duckling sign |
| 3 | Sun exposure, mole and the melanoma risk | Description of endogenous, or inherited, risk factors, and exogenous, or independent, risk factors.<br>Photo-protection and tips to ensure correct sun exposure |
| 4 | Diagnosing melanoma | Explanation of self-examination importance and of the GP and dermatologist role |
| 5 | Clinical diagnosis of melanoma | Description and iconographic documentation of the different clinical spectrum of melanoma and special clinical forms related to degree of pigmentation and/or location |
| 6 | Clinical differential diagnosis | Description and iconographic documentation of the different clinical spectrum of benign melanocytic lesions (nevi) and benign or malignant non-melanocytic lesions |
| 7 | Early diagnosis with instrumental assessment | Dermoscopy analysis and other non-invasive instrumental methods such as sequential dermoscopy, total body photography and confocal microscopy.<br>Dermoscopical and histopathological correlations. Description and iconographic documentation of the clinical and dermoscopical melanocytic nevi, melanoma and benign or malignant non-melanocytic lesions |
| 8 | Histological diagnosis | Histopathological and molecular diagnosis of melanoma and melanocytic lesions recommended by *WHO Classification of Skin Tumours* 4th edition |
| 9 | Surgery, staging, prognosis and treatment | Comprehensive review of surgical treatment, staging, prognosis, follow-up and treatment topics |
| 10 | Questions patients ask | List of the most frequently asked questions from patients, with the related answers |

### 3.3 Phases of the Study

Figure 1 illustrates an outline of the project. The project is divided into the following phases: 1) presentation of the project synopsis to the IMI centers, 2) the communication of participation in the project by the IMI centers 3) the administration of a pre-training questionnaire about the basic knowledge of the disease, the recognition and management of suspicious lesions, 4) the administration of the training course, 5) the administration of a post-training questionnaire to those participants who have filled in the pre-training questionnaire, and 6) the comparison of pre and post-training answers to evaluate the training course impact.

**Figure 1.**
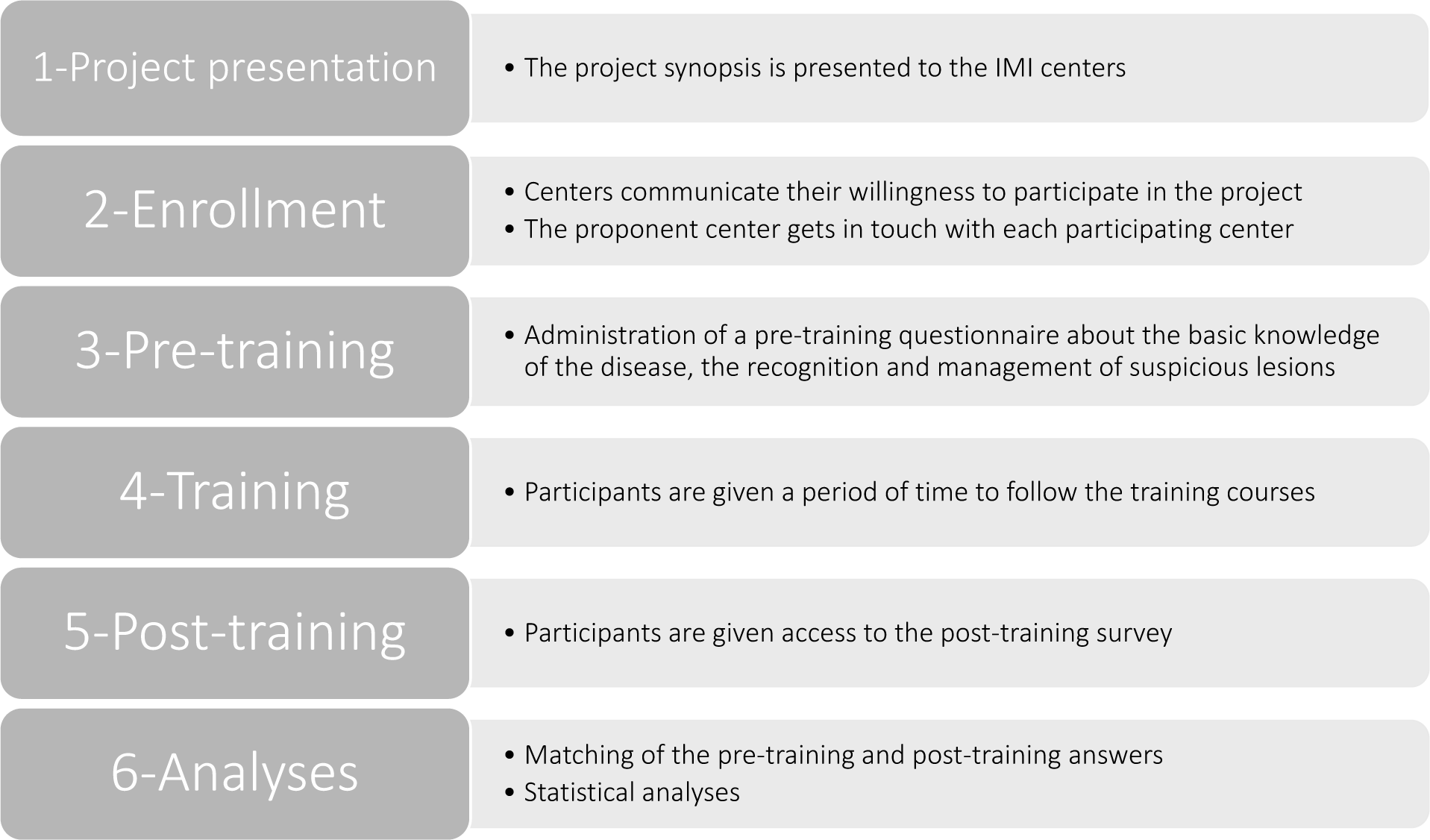
Name and description of the study phases.

### 3.4 Inclusion and exclusion criteria

All GPs, resident GPs and paediatricians of the healthcare system areas of the IMI centers are eligible. Participation is optional and, in case of participation, it is mandatory for participants to provide personal data processing consent, after reading and understanding the information on the processing of personal data according to EU Regulation 679/2016. There are no exclusion criteria.

## 4. METHODOLOGY

### 4.1 Pre-training and post-training questionnaire

The pre-training questionnaire is administered to eligible participants via Google Forms. To maximize the number of eligible people filling in the questionnaire, the coordinating centre and each participating centre arrange, in several calendar dates, a webinar to present the “MelaMEd project”, inviting in the meantime the participants to fill in the questionnaire. The link to the questionnaire should, therefore, be sent via email in advance. We have chosen Google Forms because it allows us to easily fill in the questionnaire via smartphone. Each eligible person will also receive a numerical code, required in both the pre and post-training questionnaires. This code is used to match the answers during the analysis.

The questionnaire includes 7 multiple choice theoretical questions about the prevention of skin tumours and 10 images of skin tumours to be evaluated indicating 1) whether the lesion is benign or malignant, 2) one of 4 possible diagnoses, 3) whether the patient should be referred to a specialist for a medical consult and possible excision (Supplementary Table 1). The 10 images of pigmented lesions have been selected according to two criteria: (1) they are present in the MelaMEd platform, and (2) they reflect pigmented lesions commonly observed in general practice. These criteria allow us to assume that the participants’ diagnostic skills observed in the questionnaire can be generalized to routine medical practice.

In the post-training questionnaire (Supplementary Table 1), the same questions as those included in the pre-training one will be administered, together with 10 additional questions on the Likert scale for the training course global evaluation.^20^

### 4.2 Questionnaire validation

Each question in the pre- and post-training questionnaires was extrapolated by recommendations by international publications^7,17,19,21,22^ and by clinician guideline determinants by accredited by the National Health Institute and European organizations.^3–5^ Furthermore, 20 people randomly chosen among those eligible have been asked via Google Forms for feedback about the clarity and understandability of each question. This methodology ensures that every respondent must, first of all, understand the question they are reading, strengthening the results.^23^ Questions that has been evaluated include the 7 theoretical questions and the 10 questions about the images, for a total of 17 questions. For the majority (14 out of 20) of participants, all the 17 questions (100%) were asked in a clear and understandable way. For the remaining 6 participants, there were from 1 to 3, out of 17, not clear and understandable questions (**Supplementary Table 2**). Furthermore, 9/17 questions were clear and understandable for 20 (100%) of participants, 6/17 questions for the 95%, and, lastly, 2/17 questions were clear and understandable for the 90% of participants. These figures guarantee the strength of the results, once available.

### 4.3 Consent to personal data processing

The project handles participants’ data, including name, last name, email address and workplace. An information form will be issued to participants in accordance with EU Regulation 679/2016 regarding the processing of personal data. At the beginning of the pre-training questionnaire, participants will be asked to confirm that they have read and understood the information form above mentioned and whether they authorize their data processing. Should a respondent decline authorization, the survey will instantly end, excluding the participant from the training course impact evaluation but not preventing his/her access to the asynchronous e-learning course and to the MelaMEd platform. A notice is shown prior to the questionnaire automatic closure, alerting participants in case they accidentally denied permission.

## 5. STATISTICAL ANALYSIS

Descriptive, univariate, and multivariate analysis of the questionnaire responses will be used to assess how the training program improved the participants’ diagnostic performance.

The percentages of correct answers for the theoretical questions given before and after the training will be compared.

As for the answers to the questions relating to images, we will compare the percentages of correctly diagnosed cases of benign or malignant melanoma, the percentages of right diagnoses, and the percentages of referrals to dermatologists before and after training.

With reference to the answers to the global evaluation questions, the frequency distribution will be calculated.

The Wilcoxon signed-ranks test will be used to compare the pre-training and post-training medians, and the McNemar test for paired data will be used to compare the pre-training and post-training proportions. We will use logistic regression models to investigate possible factors which might affect the accuracy of the diagnosis.

## 6. DISSEMINATION

Results will be published via (inter)national peer-reviewed journals and the findings of the study will be communicated using a comprehensive dissemination strategy aimed at healthcare professionals, in particular GPs, resident GPs and paediatricians.

## Contributions

Ignazio Stanganelli and Federica Zamagni conceived this article and drafted the manuscript. Marco Brusasco, Lauro Bucchi, Emanuele Crocetti, Stefano Falcinelli, Fabio Falcini, Sara Gandini, Serena Magi, Davide Melandri, Silvia Mancini, Patrizia Re, Rosa Vattiato revised the manuscript critically for important intellectual content. All authors read and approved the final version of the manuscript.

## Ethics approval

The study design complied with the Declaration of Helsinki ethical standards and was approved by the Ethics Committee at the Romagna Cancer Institute (ID: IRST100.37; IRST identifier codes: L4P3037, wfn.27L4). The study was approved by the Scientific Committee of the Italian Melanoma Intergroup (IMI).

## Informed Consent Statement

There are no risks associated with this study. Individual patient consent was not required as anonymized data were used.

## Data Availability Statement

The anonymized dataset used in this study is available on request from the corresponding author.

## Acknowledgments

The European Institute of Oncology, Milan, Italy is partially supported by the Italian Ministry of Health with Ricerca Corrente and 5×1,000 funds.

## Working Group

The FAD MelaMEd Working Group includes:

Maria Antonietta Pizzichetta (Aviano, Trieste), Stefania Stucci, Marco Tucci (Bari), Vincenzo De Giorgi, Daniela Massi (Firenze), Paola Ghiorzo, Cesare Massone, Enrica Tanda (Genova), Francesco de Rosa, Matelda Medri (Meldola), Roberto Patuzzo (Milano), Corrado Caracò, Marco Palla (Napoli), Alessio Fabozzi (Padova), Mario Mandalà (Perugia), Riccardo Marconcini (Pisa), Paolo Fava, Elena Marra, Pietro Quaglino, Simone Ribero (Torino) for the development of educational contents.

## Supplementary material

**Supplementary Table 1.** Question and answers of the pre-training and post-training questionnaires. The right answer is underlined.

| Question number | Question text | Multiple choice answer |
| --- | --- | --- |
| *Consent and demographic questions* |  |  |
| 1 | In accordance with EU Regulation 679/2016 regarding personal data protection, do you consent to the processing of your personal data? | a) Yes<br>b) No |
| 2 | Select your job title | a) General Practitioner<br>b) Resident General Practitioner<br>c) Paediatrician |
| 3 | What age group do you belong to? | a) <40<br>b) 40-60<br>c) >60 |
| *Theoretical questions* |  |  |
| 1* | Which of the following is a risk factor for melanoma? | a) <u>Ultraviolet radiation</u><br>b) Cigarette smoke<br>c) Consumption of red meat<br>d) Alcohol abuse |
| 2* | What is needed for a complete visual examination of the skin? | a) Evaluation only of the suspected neoformation reported by the patient<br>b) Live evaluation of the entire skin area in dim light<br>c) <u>Live evaluation of the entire skin area in optimal lighting conditions and with the use of a magnifying glass</u><br>d) Referral to the specialist reference centre |
| 3* | What does the ABCDE acronym for the recognition of a suspected melanoma mean? | a) Asymmetry, Regular edges, Homogeneous colour, Dimensions <2 mm, Empathy<br>b) Asymmetry, Irregular edges, Homogeneous colour, Dimensions <5 mm, Static evolution<br>c) <u>Asymmetry, Irregular edges, Uneven colour, Dimensions >6 mm, rapid Evolution</u><br>d) Asymmetry, Irregular edges, Homogeneous color, Small dimensions, Entropy<br>e) I don't know the ABCDE acronym |
| 4* | What does the EFG rule coined by Australian dermatologists for the recognition of suspected nodular melanoma mean? | a) Erythema, Firm, Ground<br>b) <u>Elevation, Firm, Growth</u><br>c) Evolution, Figure, Great dimension<br>d) Extension, Family history, Great dimension<br>e) I don't know the EFG rule |
| 5* | What is meant by the "ugly duckling" sign for recognizing a suspected melanoma? | a) Homogeneous nevus equal to other nevus in shape and color<br>b) Nevus regular compared to other nevus<br>c) Moriform congenital nevus<br>d) <u>Different nevus that "stands out" compared to other nevus due to its shape, color and/or rapid evolution</u> |
|  |  | e) I don't know the sign of the "ugly duckling" |
| 6* | Which of the following statements about dermoscopy is true? | a) <u>Dermoscopy is a non-invasive technique that allows the in vivo evaluation of skin colors and structures not visible to the naked eye</u><br>b) Dermoscopy is a non-invasive technique that uses a laser beam with a near-infrared wavelength (830 nm), to evaluate the cells of the superficial layer of the skin at a depth of approximately 100-200 µm<br>c) Dermoscopy is an incisional biopsy specific for melanocytic lesions<br>d) Dermoscopy can replace histological examination |
| 7* | Which of the following statements about Breslow thickness is true? | a) It indicates the degree of invasion of the skin. It is divided into 5 levels based on the degree of infiltration: from I (melanoma in situ) to V (infiltration of the entire dermis up to the adipose tissue)<br>b) <u>It indicates the thickness of the melanoma from the granular layer of the skin (or from the base of the ulcer if the lesion is ulcerated) to the point of maximum infiltration into the dermis. It is measured in mm</u><br>c) It indicates whether the melanoma is in the horizontal or vertical histological phase<br>d) It indicates the depth of tumor-infiltrating lymphocytes (TILs) |
| *Images* |  |  |
| 1* | Consider the following pigmented lesion, indicate if it is: | a) Benign<br>b) Malignant<br>c) I don't know |
|  | Would you refer the patient to a dermatologist? | a) Yes<br>b) No |
|  | Which is your diagnosis? | a) Common Nevus<br>b) Atypical Nevus<br>c) <u>Melanoma</u><br>d) Seborrheic Keratosis |
| 2* | Consider the following pigmented lesion, indicate if it is: | a) Benign<br>b) Malignant<br>c) I don't know |
|  | Would you refer the patient to a dermatologist? | a) Yes<br>b) No |
|  | Which is your diagnosis? | a) Seborrheic Keratosis<br>b) Common Nevus<br>c) <u>Congenital Nevus</u><br>d) Melanoma |
| 3* | Consider the following pigmented lesion, indicate if it is: | a) Benign<br>b) Malignant<br>c) I don't know |
|  | Would you refer the patient to a dermatologist? | a) Yes<br>b) No |
|  | Which is your diagnosis? | a) <u>Seborrheic Keratosis</u><br>b) Common Nevus<br>c) Atypical Nevus<br>d) Melanoma |
| 4* | Consider the following pigmented lesion, indicate if it is: | a) Benign<br>b) Malignant<br>c) I don't know |
|  | Would you refer the patient to a dermatologist? | a) Yes<br>b) No |
|  | Which is your diagnosis? | a) Seborrheic Keratosis<br>b) Common Nevus<br>c) Congenital Nevus<br>d) <u>Melanoma</u> |
| 5* | Consider the following pigmented lesion, indicate if it is: | a) Benign<br>b) Malignant<br>c) I don't know |
|  | Would you refer the patient to a dermatologist? | a) Yes<br>b) No |
|  | Which is your diagnosis? | a) Congenital Nevus<br>b) <u>Melanoma</u><br>c) Seborrheic Keratosis<br>d) Basal cell carcinoma |
| 6* | Consider the following pigmented lesion, indicate if it is: | a) Benign<br>b) Malignant<br>c) I don't know |
|  | Would you refer the patient to a dermatologist? | a) Yes<br>b) No |
|  | Which is your diagnosis? | a) <u>Melanocytic Nevus</u><br>b) Basal cell carcinoma<br>c) Melanoma<br>d) Seborrheic Keratosis |
| 7* | Consider the following pigmented lesion, indicate if it is: | a) Benign<br>b) Malignant<br>c) I don't know |
|  | Would you refer the patient to a dermatologist? | a) Yes<br>b) No |
|  | Which is your diagnosis? | a) Atypical Nevus<br>b) <u>Congenital Nevus</u><br>c) Melanoma<br>a) Basal cell carcinoma |
| 8* | Consider the following pigmented lesion, indicate if it is: | a) Benign<br>b) Malignant<br>c) I don't know |
|  | Would you refer the patient to a dermatologist? | a) Yes<br>b) No |
|  | Which is your diagnosis? | a) Melanocytic Nevus<br>b) Angioma<br>c) <u>Melanoma</u><br>d) Basal Cell Carcinoma |
| 9* | Consider the following pigmented lesion, indicate if it is: | a) Benign<br>b) Malignant<br>c) I don't know |
|  | Would you refer the patient to a dermatologist? | a) Yes<br>b) No |
|  | Which is your diagnosis? | a) Solar Lentigo<br>b) Seborrheic Keratosis<br>c) Congenital Nevus<br>d) <u>Malignant Lentigo</u> |
| 10* | Consider the following pigmented lesion, indicate if it is: | a) Benign<br>b) Malignant<br>c) I don't know |
|  | Would you refer the patient to a dermatologist? | a) Yes<br>b) No |
|  | Which is your diagnosis? | a) Seborrheic Keratosis<br>b) <u>Basal Cell Carcinoma</u><br>c) Congenital Nevus<br>d) Melanoma |
| *Global evaluation questions* |  |  |
| 1* | How confident do you feel in giving patients information on the primary prevention of skin cancer? | a) Absolutely not confident<br>b) Not confident<br>c) Don't know<br>d) Confident<br>e) Absolutely confident |
| 2* | How confident do you feel in informing the patient about the skin self-examination? | a) Absolutely not confident<br>b) Not confident<br>c) Don't know<br>d) Confident<br>e) Absolutely confident |
| 3* | How confident do you feel in making a diagnosis of a suspected melanoma? | a) Absolutely not confident<br>b) Not confident<br>c) Don't know<br>d) Confident<br>e) Absolutely confident |
| 4** | Do you think MelaMEd has improved your ability to recognize patients at high risk of melanoma? | a) Absolutely disagree<br>b) Disagree<br>c) Don't know<br>d) Agree<br>e) Absolutely agree |
| 5** | Do you think MelaMEd has improved your ability to distinguish melanoma from benign lesions? | a) Absolutely disagree<br>b) Disagree<br>c) Don't know<br>d) Agree<br>e) Absolutely agree |
| 6** | Do you think MelaMEd has improved your knowledge in skin cancer management? | a) Absolutely disagree<br>b) Disagree<br>c) Don't know<br>d) Agree<br>e) Absolutely agree |
| 7** | Do you prefer multimedia training based on the MelaMEd model to traditional live training? | a) Absolutely disagree<br>b) Disagree<br>c) Don't know |
|  |  | d) Agree<br>e) Absolutely agree |
| 8** | Do you think that the active participation of GPs, resident GPs and paediatricians can improve the early diagnosis of melanoma? | a) Absolutely disagree<br>b) Disagree<br>c) Don't know<br>d) Agree<br>e) Absolutely agree |
| 9** | Would you be interested in attending a dermoscopy course? | a) Yes<br>b) No |
| 10** | Would you like to receive additional information about melanoma on a regular basis? | a) Yes<br>b) No |

GP, General practitioner;
\*, question present both in pre-training and post-training questionnaire
\*\*, question present only in the post-training questionnaire

**Supplementary Table 2.** Distribution of the number of clear and understandable questions from the pre and post-training questionnaire. Questions that has been evaluated include the 7 theoretical questions and the 10 questions about the images, for a total of 17 questions.

| Participants | Clear and understandable questions<br>N(%) | Not clear and understandable questions<br>N(%) |
| --- | --- | --- |
| 14 | 17 (100) | 0 (0) |
| 3 | 16 (94) | 1 (6) |
| 2 | 15 (88) | 2 (12) |
| 1 | 14 (82) | 3 (18) |

